# Associations of the 2018 World Cancer Research Fund/American Institute of Cancer Research (WCRF/AICR) Cancer Prevention Recommendations with Stages of Colorectal Carcinogenesis

**DOI:** 10.1101/2023.01.24.23284936

**Authors:** Ane Sørlie Kværner, Astrid Riseth Andersen, Hege Berg Henriksen, Markus Knudsen, Anne Marte Wetting Johansen, Anette Hjartåker, Siv Kjølsrud Bøhn, Ingvild Paur, Gro Wiedswang, Sigbjørn Smeland, Trine B. Rounge, Rune Blomhoff, Paula Berstad

## Abstract

While adherence to cancer prevention recommendations is linked to lower risk of colorectal cancer (CRC), few have studied associations across the entire spectrum of colorectal carcinogenesis. Here, we studied the relationship of the standardized 2018 World Cancer Research Fund/American Institute for Cancer Research (WCRF/AICR) Score for cancer prevention recommendations with colorectal carcinogenesis in a cross-sectional setting. Baseline data from two studies was combined to measure adherence to the seven-point 2018 WCRF/AICR Score in screening participants with a positive faecal immunochemical test and CRC patients in an intervention study. Dietary intake, body fatness and physical activity were assessed using self-administered questionnaires. Multinomial logistic regression was used to estimate odds ratios (ORs) and 95% confidence intervals (CIs) for screen-detected colorectal lesions and CRC. Of 1,914 participants, 548 were free from adenomas, 524 had non-advanced adenomas, 349 had advances lesions and 493 had CRC (63 screen-detected and 430 recruited from the intervention study). Adherence to the 2018 WCRF/AICR Score was inversely associated with advanced colorectal lesions; OR 0.82 (95% CI 0.71, 0.94, *p*_*trend*_ 0.005) per score point, but not CRC. Adherence to the alcohol recommendation was the single factor most strongly inversely associated with CRC development, being significantly associated with advanced colorectal lesions and CRC. Adherence to the 2018 WCRF/AICR cancer prevention recommendations was associated with lower probability of screen-detected advanced colorectal lesions, but not CRC. Taking a holistic approach to cancer prevention is important to prevent the occurrence of precancerous colorectal lesions.

**What’s new?:** While several studies have documented an association between adherence to cancer prevention recommendations and risk colorectal cancer, data is sparse when it comes to the precancerous lesions. In this study, including participants representing the entire spectrum of colorectal carcinogenesis, strong inverse associations were observed between adherence to the 2018 World Cancer Research Fund/American Institute of Cancer Research (WCRF/AICR) and the two main precursor lesion types (advanced adenoma and advanced serrated lesion), highlighting the importance of adopting a healthy lifestyle early on to prevent the development of colorectal cancer.

**Trial Registration:** ClinicalTrials.gov Identifier: NCT01538550 (Bowel Cancer Screening in Norway (BCSN) trial) and NCT01570010 (CRC-NORDIET).

## Introduction

Globally, colorectal cancer (CRC) is the third most diagnosed cancer in women and men, accounting for over 1.9 million incident cases and 900.000 deaths in 2020 (1). It has been estimated that about half of all CRC cases could have been avoided by following a healthy diet, being physically active and maintaining a healthy body weight (2). In 2018, the World Cancer Research Fund (WCRF) and American Institute for Cancer Research (AICR) issued an expert report (3^rd^ edition since 1997) summarizing the evidence on risk and preventing factors of CRC and other common cancers (2). The report concluded with a list of ten recommendations concerning body weight, physical activity, diet and breastfeeding aiming at reducing cancer risk and improve overall cancer survival. A standardized scoring system (“the 2018 WCRF/AICR Score”) has been developed to measure adherence to eight of these ten recommendations (3,4). While there is strong evidence that adherence to select components of the WCRF/AICR Score (e.g. maintaining a healthy body weight and being physically active) protect against CRC (2), few studies have examined the joint effect of these on colorectal carcinogenesis, especially for early stage disease development. To the best of our knowledge, no previous study has examined adherence to the 2018 WCRF/AICR Score across the entire spectrum of colorectal carcinogenesis. We therefore investigated the associations of adherence to the standardized 2018 WCRF/AICR Score (excluding the component on breast feeding) with occurrence of colorectal lesions at various stages of the carcinogenic process. We also examined associations for the seven individual components of the score.

## Methods

### The BCSN and the CRCbiome study

The CRCbiome study is a prospective cohort sub study within the Bowel Cancer Screening in Norway (BCSN) trial (5), with the overall aim of developing a microbiome-based classifier for improved detection of advanced colorectal lesions at screening (6). BCSN participants, aged 50-74 years in 2012, who had tested positive for an immunochemical fecal occult blood test (FIT) during 2017-2021, were eligible for the study. Hemoglobin levels above 15 μg/g feces were considered positive and qualified for colonoscopy referral. Participants were invited to the study after being informed about their test result, but before attending follow-up colonoscopy in one of the two screening centres in Moss and Bærum hospitals in South-East Norway. With the invitation letter, participants received two questionnaires to be completed prior to colonoscopy: a lifestyle- and demographics questionnaire (LDQ) and a food frequency questionnaire (FFQ). Returning at least one of the questionnaires was regarded as consent to the study. Of 2,698 invited, 1,653 (61%) agreed to participate.

The BCSN and the CRCbiome study have been approved by the Regional Committee for Medical Research Ethics in South East Norway (Approval no.: 2011/1272 and 63148, respectively). The BCSN is also registered at clinicaltrials.gov (National clinical trial (NCT) no.: 01538550).

### The CRC-NORDIET study

The CRC-NORDIET study is a randomized controlled trial with two parallel study arms (only baseline data used in the current study). The overall aim is to investigate the effect of a diet in accordance with the Norwegian food based dietary guidelines on disease-free survival and overall survival among Norwegian CRC patients (7). The participants were recruited between 2012 and 2020. Women and men aged 50-80 years diagnosed with primary invasive CRC at Akershus or Oslo University Hospital were eligible for the study. The cancer needed to be an established primary adenocarcinoma in the colon or the rectum and classified by the ICD-codes C18-20, with TNM stages I-III. Participants were invited at the hospitals prior to surgery or by telephone after surgery, and an informed consent needed to be signed before randomization to either the intervention group or the control group. Of 621 participants signing the consent, 503 were eligible for the study and participated at baseline. The CRC-NORDIET study has been approved by the Regional Committee for Medical Research Ethics in South East Norway (Approval no.:2011/836). It is also registered at clinicaltrials.gov (NCT no.: 01570010).

### Study sample

In the current study, participants with available dietary information by October 2021 from the CRCbiome (n=1,616) and the CRC-NORDIET (n=464) study were included. Exclusion criteria included non-attendance on the follow-up colonoscopy in CRCbiome (n=39) or withdrawal from the study after inclusion (n=15), presence of a metastatic disease (n=4 in total), delivery of a poor quality FFQ (n=21 from CRCbiome) or reporting a too low (<600 and <800 kcal/day for women and men, respectively, n=9 in total) or too high (>3,500 and >4,200 kcal/day for women and men, respectively, n=78 in total) energy intake (**Figure 1**). This resulted in a study population of 1,914 participants, 1,484 from CRCbiome and 430 from CRC-NORDIET. Of the participants in CRCbiome, 548 were free from any adenomas, 524 had one or more non-advanced adenomas, 394 had one or more advanced lesions and 63 had a non-metastatic CRC.

**Figure 1.**
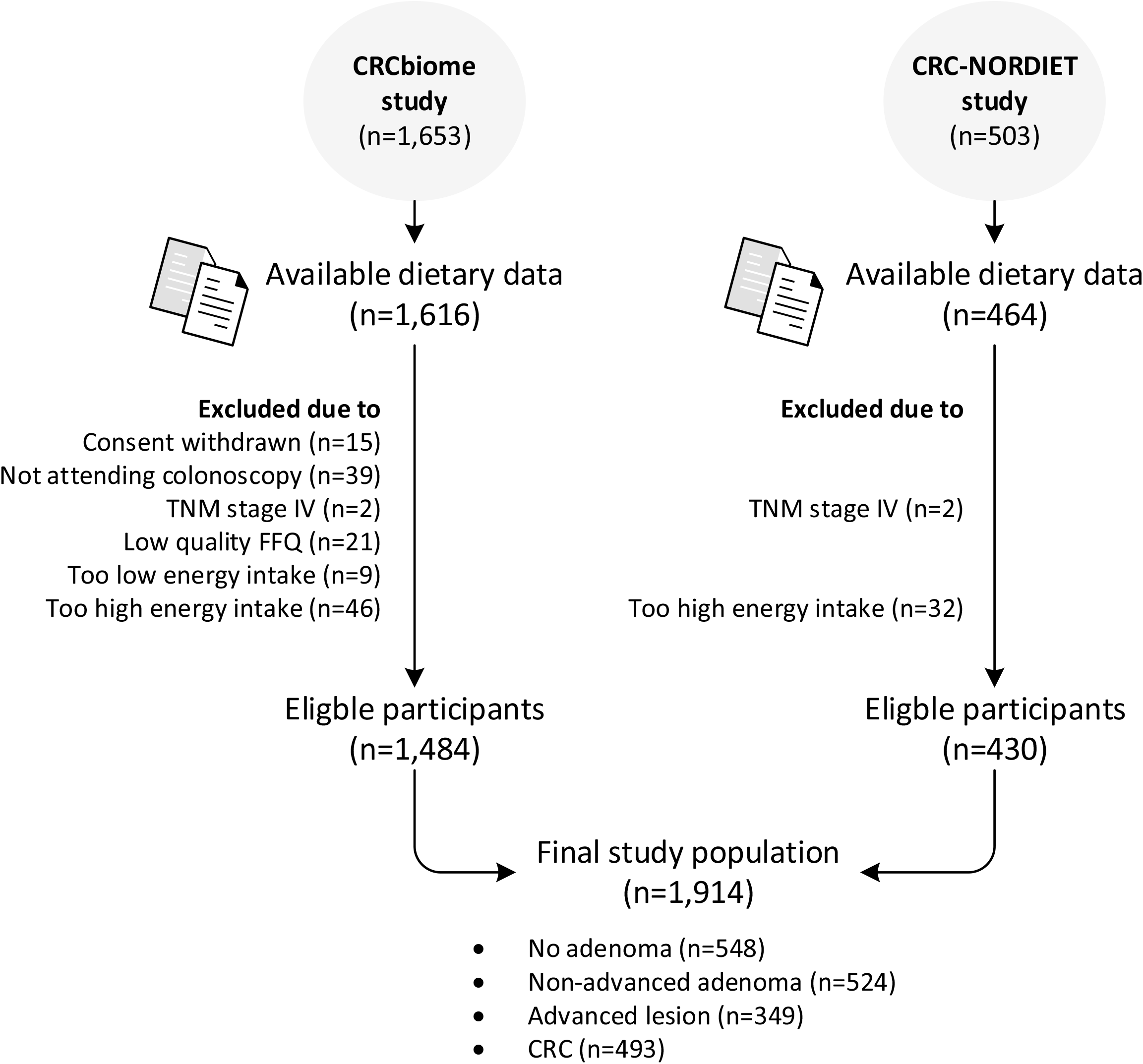
Flowchart of the study participants.

### Assessment of dietary intake, body fatness and physical activity

In both studies, dietary data were obtained using self-administered semiquantitative FFQs, designed to capture the habitual diet the preceding year. The two questionnaires are modified versions of an FFQ developed by the Department of Nutrition, University of Oslo, which has been validated for a variety of nutrients and food groups (8–10). The version used in CRCbiome had 23 main questions covering 256 food items and an open field for entries not covered by the questionnaire. The CRC-NORDIET version had 24 main questions covering 282 food items and an open field for entries not covered by the questionnaire. For each food item, participants were asked to record frequency of consumption, ranging from never/seldom to several times a day, and/or amount, typically as portion size given in various household units. Dietary intake was calculated using the food and nutrient calculation system, ‘kostberegningssystem’ (KBS), and the associated database AE-18, developed at the Department of Nutrition, University of Oslo. AE-18 is an extended version of the official Norwegian Food Composition Table (11). Prior to analyses, all questionnaires were reviewed and evaluated by trained personnel according to “Tutorial for scanning of FFQs and food diaries” prepared by the Department of Nutrition, University of Oslo. Body mass index was calculated from self-reported body height and weight in the FFQ in the CRCbiome and the CRC-NORDIET study. Physical activity level was covered by questions on total number of weekly hours of activity on three different intensity level in the lifestyle and demographic questionnaire (LDQ) in the CRCbiome study. In CRC-NORDIET, data on physical activity was obtained from a short semi-quantitative validated FFQ called NORDIET-FFQ (12,13).

### Operationalization of the 2018 WCRF/AICR Score

The 2018 WCRF/AICR recommendations for cancer prevention were operationalized following a standardized scoring system developed by Shams-White, *et al*. in 2019 (3,4). Eight of the ten cancer prevention recommendations were included: 1) be a healthy weight, 2) be physically active, 3) consume a diet rich in wholegrains, vegetables, fruit and beans, 4) limit consumption of “fast foods” and other processed foods high in fat, starches or sugars, 5) limit consumption of red and processed meat, 6) limit consumption of sugar sweetened drinks, 7) limit alcohol consumption and 8) for mothers, breastfeed your baby, if you can (optional). The two remaining recommendations, i.e. ‘do not use supplements’ and ‘after a cancer diagnosis, follow our recommendations, if you can’ were left out due to operational redundancy. In the present study, all recommendations were included, except the one concerning breastfeeding due to lack of data. For each recommendation, participants could earn 1, 0.5 or 0 points for fully, partially and not meeting the recommendation, respectively. The total score therefore ranged from 0 to 7 points, higher scores indicating greater adherence to the recommendations. To promote transparency and reproducibility, a detailed overview of how each recommendation was operationalized is given in **Supplementary Table 1**, as encouraged by the developers of the score (4).

**Table 1.** Key characteristics of the study population by study and disease stage (n=1,914)^1^.

| Variables | CRCbiome<br>(n=1,484) |  |  |  | CRC-NORDIET<br>(n=430) |
| --- | --- | --- | --- | --- | --- |
|  | No adenoma<br>(n=548) | Non-advanced<br>adenoma<br>(n=524) | Advanced lesion<br>(n=349) | CRC<br>(n=63) | CRC<br>(n=430) |
| **Demography and lifestyle** |  |  |  |  |  |
| Age, years | 65.8 (60.7, 71.0) | 67.8 (62.8, 72.5) | 67.7 (62.5, 72.0) | 68.0 (62.5, 72.8) | 67.0 (60.0, 72.0) |
| Male sex, *n* (%) | 267 (48.7) | 305 (58.2) | 218 (62.5) | 35 (55.6) | 232 (54.0) |
| Nationality, *n* (%) |  |  |  |  |  |
| Native | 501 (91.4) | 471 (89.9) | 316 (90.5) | 56 (88.9) | 303 (70.5) |
| Non-native | 34 (6.2) | 27 (5.1) | 16 (4.6) | 5 (7.9) | 17 (4.0) |
| Missing | 13 (2.4) | 26 (5.0) | 17 (4.9) | 2 (3.2) | 110 (25.6) |
| Family history of CRC, *n* (%) |  |  |  |  |  |
| Yes | 84 (15.3) | 89 (17.0) | 65 (18.6) | 17 (27.0) | 65 (15.1) |
| No | 407 (74.3) | 390 (74.4) | 252 (72.2) | 41 (65.1) | 229 (53.3) |
| Unknown | 57 (10.4) | 45 (8.6) | 32 (9.2) | 5 (7.9) | 136 (31.6) |
| Education, *n* (%) |  |  |  |  |  |
| Primary school | 93 (17.0) | 91 (17.4) | 58 (16.6) | 9 (14.3) | 42 (9.8) |
| High school | 223 (40.7) | 195 (37.2) | 134 (38.4) | 27 (42.9) | 177 (41.2) |
| University/college | 225 (41.1) | 231 (44.1) | 147 (42.1) | 27 (42.9) | 202 (47.0) |
| Missing | 7 (1.3) | 7 (1.3) | 10 (2.9) | 0 (0.0) | 9 (2.1) |
| Marital status, *n* (%) |  |  |  |  |  |
| Married/cohabiting | 454 (82.8) | 399 (76.1) | 267 (76.5) | 50 (79.4) | 305 (70.9) |
| Not married/non-cohabiting | 89 (16.2) | 119 (22.7) | 72 (20.6) | 13 (20.6) | 116 (27.0) |
| Missing | 5 (0.9) | 6 (1.1) | 10 (2.9) | 0 (0.0) | 9 (2.1) |
| Working status, *n* (%) |  |  |  |  |  |
| Employed | 199 (36.3) | 171 (32.6) | 109 (31.2) | 18 (28.6) | 121 (28.1) |
| Retired/unemployed | 343 (62.6) | 347 (66.2) | 230 (65.9) | 45 (71.4) | 295 (68.6) |
| Missing | 6 (1.1) | 6 (1.1) | 10 (2.9) | 0 (0.0) | 14 (3.3) |
| Smoking status, *n* (%) |  |  |  |  |  |
| Current smoker | 63 (11.5) | 86 (16.4) | 59 (16.9) | 6 (9.5) | 44 (10.2) |
| Non smoker | 478 (87.2) | 430 (82.1) | 281 (80.5) | 57 (90.5) | 385 (89.5) |
| Missing | 7 (1.3) | 8 (1.5) | 9 (2.6) | 0 (0.0) | 1 (0.2) |
| **Clinical information** |  |  |  |  |  |
| Hospital, *n* (%) |  |  |  |  |  |
| Bærum | 237 (43.2) | 261 (49.8) | 179 (51.3) | 34 (54.0) | - |
| Moss | 311 (56.8) | 263 (50.2) | 170 (48.7) | 29 (46.0) | - |
| Ullevål | - | - | - | - | 221 (51.4) |
| Akershus | - | - | - | - | 209 (48.6) |
| Tumor localization<sup>2</sup>, *n* (%) |  |  |  |  |  |
| Colon | - | - | - | 32 (50.8) | 251 (58.4) |
| Rectum | - | - | - | 31 (49.2) | 174 (40.5) |
| Missing | - | - | - | 0 (0.0) | 5 (1.2) |
| TNM stage, *n* (%) |  |  |  |  |  |
| I | - | - | - | 36 (57.1) | 115 (26.7) |
| II | - | - | - | 17 (27.0) | 143 (33.3) |
| III | - | - | - | 10 (15.9) | 127 (29.5) |
| Missing | - | - | - | 0 (0.0) | 45 (10.5) |
| **Completion of the FFQ** |  |  |  |  |  |
| Diagnosis known, *n* (%) |  |  |  |  |  |
| Yes | 31 (5.7) | 56 (10.7) | 36 (10.3) | 3 (4.8) | 430 (100) |
| No | 517 (94.3) | 468 (89.3) | 313 (89.7) | 60 (95.2) | 0 (0.0) |
| Time relative to hospital admission, days | -7 (-13, -2) | -5.5 (-13, -1) | -5 (-14, -1) | -8 (-13.5, -3) | 86 (-0.5, 127) |

<sup>1</sup>Values are median (p25, p75) for continuous variables and n (%) for categorical variables.
<sup>2</sup>One of the colon cancer cases in CRCbiome was also diagnosed with a primary invasive rectum cancer.
Abbreviations: CRC; colorectal cancer, FFQ; food frequency questionnaire, n; number, TNM; Tumor, node, metastasis.

### Assessment of covariates

Demographic data (i.e. level of education, working status, nationality and marital status) and information on smoking status were retrieved from the LDQ in the CRCbiome study. The LDQ is a self-administered, four-page questionnaire which was piloted in a targeted population prior to study start and adjusted according to participants’ feedback. In the CRC-NORDIET study, demography variables were collected from a self-administered, one-page questionnaire. Smoking status was reported in the previously mentioned FFQ. Further details regarding the studies have been described previously (6,7). The data sets from the two studies were combined and harmonized to ensure comparability prior to analyses.

### Outcome assessment

In the CRCbiome study, the follow-up colonoscopy formed the basis for the outcome classification. Presence and clinicopathological characteristics of detected lesions were registered by the responsible gastroenterologist using a structured recording system. In the CRC-NORDIET study – where all participants were recruited based on their cancer diagnosis, tumor characteristics, including disease severity and localization, were retrieved from electronic patient records.

Based on the available clinicopathological information in the two studies, participants were categorized into the following diagnostic groups: No adenoma, non-advanced adenoma, advanced colorectal lesions and CRC (any adenocarcinoma of the colon and rectum, i.e. ICD-10 codes C18-20). Advanced colorectal lesions included both advanced adenomas (any adenoma with villous histology, high-grade dysplasia or polyp size greater than or equal to 10 mm) and advanced serrated lesions (any serrated lesion with size ≥10 mm or dysplasia) (14). In cases of multiple findings, the most severe finding was selected.

### Statistical analyses

Descriptive statistics are given as median (p25, p75) and numbers (percentages) for continuous and categorical variables, respectively. To study correlations between the 2018 WCRF/AICR Score, including the individual diet and lifestyle components and total energy intake, scatter plots with linear trend lines were created. The proportion of participants who fully, partly and did not adhere to the recommendations by stage of the carcinogenic process was illustrated by bar charts. Multinomial logistic regression analyses were used to calculate the odds ratios (ORs) and 95% confidence intervals (CIs) for colorectal lesions (presence of non-advanced adenoma, advanced lesions and CRC) relative to no adenoma by adherence to the 2018 WCRF/AICR Score and the seven individual recommendations.

Based on the distribution of adherence to the 2018 WCRF/AICR Score, participants were divided into four exposure groups: ≤2.5 (reference category), >2.5-3.5, >3.5-4.5 and >4.5 points. Adherence was also examined on a continuous scale. For the individual diet and lifestyle recommendations, participants were divided into those fully (1 point), partially (0.5 point) and not adhering (0 points) to the recommendation, the latter being used as reference category. The main association analyses were conducted in the study population as a whole and stratified by sex. A separate analysis was also conducted by type of precursor lesion (i.e. advanced adenomas, advanced serrated lesions or the combination of the two).

For the associations between adherence to the 2018 WCRF/AICR Score and stages of the carcinogenic process, models were adjusted for the following covariates: age (continuous), sex, energy intake (continuous), smoking status (current smoker, non-smoker, missing), education level (primary school, high school, collage/university, missing) and family history of CRC (yes, no, unknown). For the individual diet and lifestyle recommendations – where the groups compared were less balanced, models were adjusted for age (continuous) and sex. The covariate selection was based on a priori knowledge on the relationship between diet and lifestyle and colorectal carcinogenesis (2,15,16).

As sensitivity analyses, we examined associations by severity of the cancer disease (TNM stage I *vs*. II and III), as well as timing of cancer care for which the diet and lifestyle registration was performed: not yet diagnosed (n=60), admitted to the hospital for CRC surgery (n=107) and in recovery (n=320). We also performed a sensitivity analyses, restricting the study sample to those recruited from the CRCbiome study only (**Supplementary Table 2**).

**Table 2.**
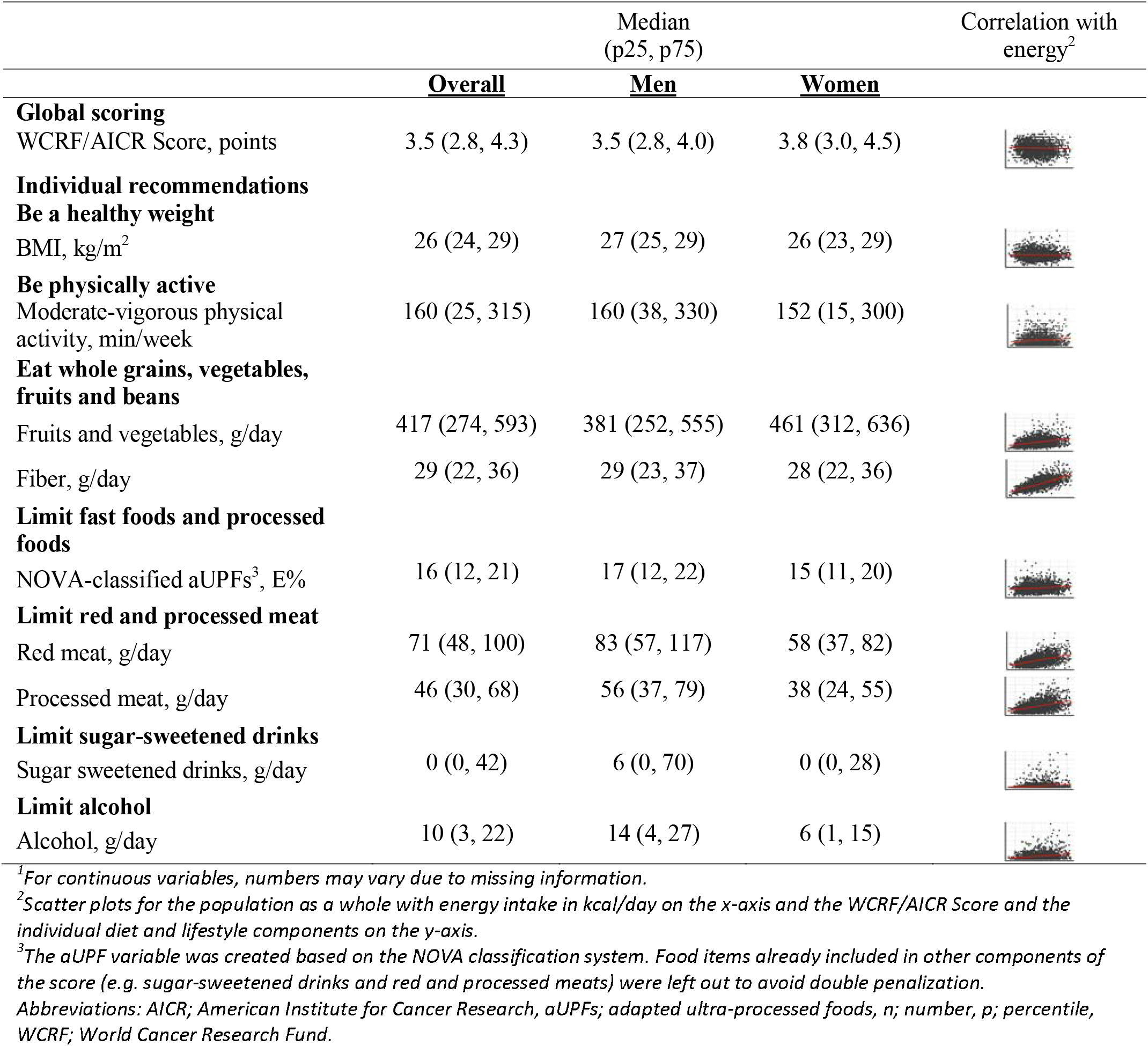
Summary of diet and lifestyle characteristics of the 2018 WCRF/AICR Score in the study population as a whole (n=1,914) and by sex (men: 1,057, women: 857)^1^.

|  |  | Median<br>(p25, p75) |  | Correlation with<br>energy<sup>2</sup> |
| --- | --- | --- | --- | --- |
|  | <u>Overall</u> | <u>Men</u> | <u>Women</u> |  |
| **Global scoring** |  |  |  |  |
| WCRF/AICR Score, points | 3.5 (2.8, 4.3) | 3.5 (2.8, 4.0) | 3.8 (3.0, 4.5) |  |
| **Individual recommendations** |  |  |  |  |
| **Be a healthy weight** |  |  |  |  |
| BMI, kg/m<sup>2</sup> | 26 (24, 29) | 27 (25, 29) | 26 (23, 29) |  |
| **Be physically active** |  |  |  |  |
| Moderate-vigorous physical activity, min/week | 160 (25, 315) | 160 (38, 330) | 152 (15, 300) |  |
| **Eat whole grains, vegetables, fruits and beans** |  |  |  |  |
| Fruits and vegetables, g/day | 417 (274, 593) | 381 (252, 555) | 461 (312, 636) |  |
| Fiber, g/day | 29 (22, 36) | 29 (23, 37) | 28 (22, 36) |  |
| **Limit fast foods and processed foods** |  |  |  |  |
| NOVA-classified aUPFs<sup>3</sup>, E% | 16 (12, 21) | 17 (12, 22) | 15 (11, 20) |  |
| **Limit red and processed meat** |  |  |  |  |
| Red meat, g/day | 71 (48, 100) | 83 (57, 117) | 58 (37, 82) |  |
| Processed meat, g/day | 46 (30, 68) | 56 (37, 79) | 38 (24, 55) |  |
| **Limit sugar-sweetened drinks** |  |  |  |  |
| Sugar sweetened drinks, g/day | 0 (0, 42) | 6 (0, 70) | 0 (0, 28) |  |
| **Limit alcohol** |  |  |  |  |
| Alcohol, g/day | 10 (3, 22) | 14 (4, 27) | 6 (1, 15) |  |

<sup>1</sup>For continuous variables, numbers may vary due to missing information.
<sup>2</sup>Scatter plots for the population as a whole with energy intake in kcal/day on the x-axis and the WCRF/AICR Score and the individual diet and lifestyle components on the y-axis.
<sup>3</sup>The aUPF variable was created based on the NOVA classification system. Food items already included in other components of the score (e.g. sugar-sweetened drinks and red and processed meats) were left out to avoid double penalization.
Abbreviations: AICR; American Institute for Cancer Research, aUPFs; adapted ultra-processed foods, n; number, p; percentile, WCRF; World Cancer Research Fund.

In line with the most recent statement from the American Statistical Association on p-values (17), emphasis was put on effect sizes, variation and uncertainty of the data rather than p-values in the interpretation of the results. All statistical analyses were performed using RStudio, version 3.6.3 (The R Foundation for Statistical Computing, Vienna, Austria). The main R packages used included those within the Tidyverse (18), as well as skimr, nnet and vgam.

## Results

### Key characteristics of the study population

Characteristics of the study population by stage of the carcinogenic process are presented in **Table 1**. The median age of participants was 67 years, ranging from 66 to 68 years across the diagnostic groups. For all carcinogenic stages, there was a dominance of male participants (54-63%). Of participants diagnosed with CRC, the screen-detected were more likely to be early-stage cancers than those recruited through the intervention study (84 contra 60% with TNM stage I and II).

### Adherence to the 2018 WCRF/AICR Score

Descriptive statistics of the 2018 WCRF/AICR Score, as well as the individual diet and lifestyle components forming the basis for the score, is provided in **Table 2**. The median (p25, p75) adherence to the recommendations was 3.5 (2.8, 4.3) points. Women scored slightly higher than men; 3.8 (3.0, 4.5) *vs*. 3.5 (2.8, 4.0) points, respectively. None of the participants adhered to all recommendations, the highest score being 6.5, achieved by 3 (0.2%) participants. For the individual components, highest adherence was seen for limiting the amounts of sugar-sweetened beverages (52% fully adhering), being physically active (49% fully adhering) and eating a diet rich in wholegrains, vegetables, fruit and beans (of which 53 and 45% fully adhered to the recommendations concerning daily fruit and vegetable intake and fiber intake, respectively). For the recommendations on having a healthy body weight and limiting the consumption of “fast foods” and other processed foods high in fat, starches and sugars, approximately one third of the participants fully adhered to the recommendations. The lowest adherence to the recommendations were seen for alcohol intake and consumption of red and processed meat, where 12% and 2% fully adhered to the recommendation, respectively. Adherence by clinical group is presented in **Figure 2**.

**Figure 2.**
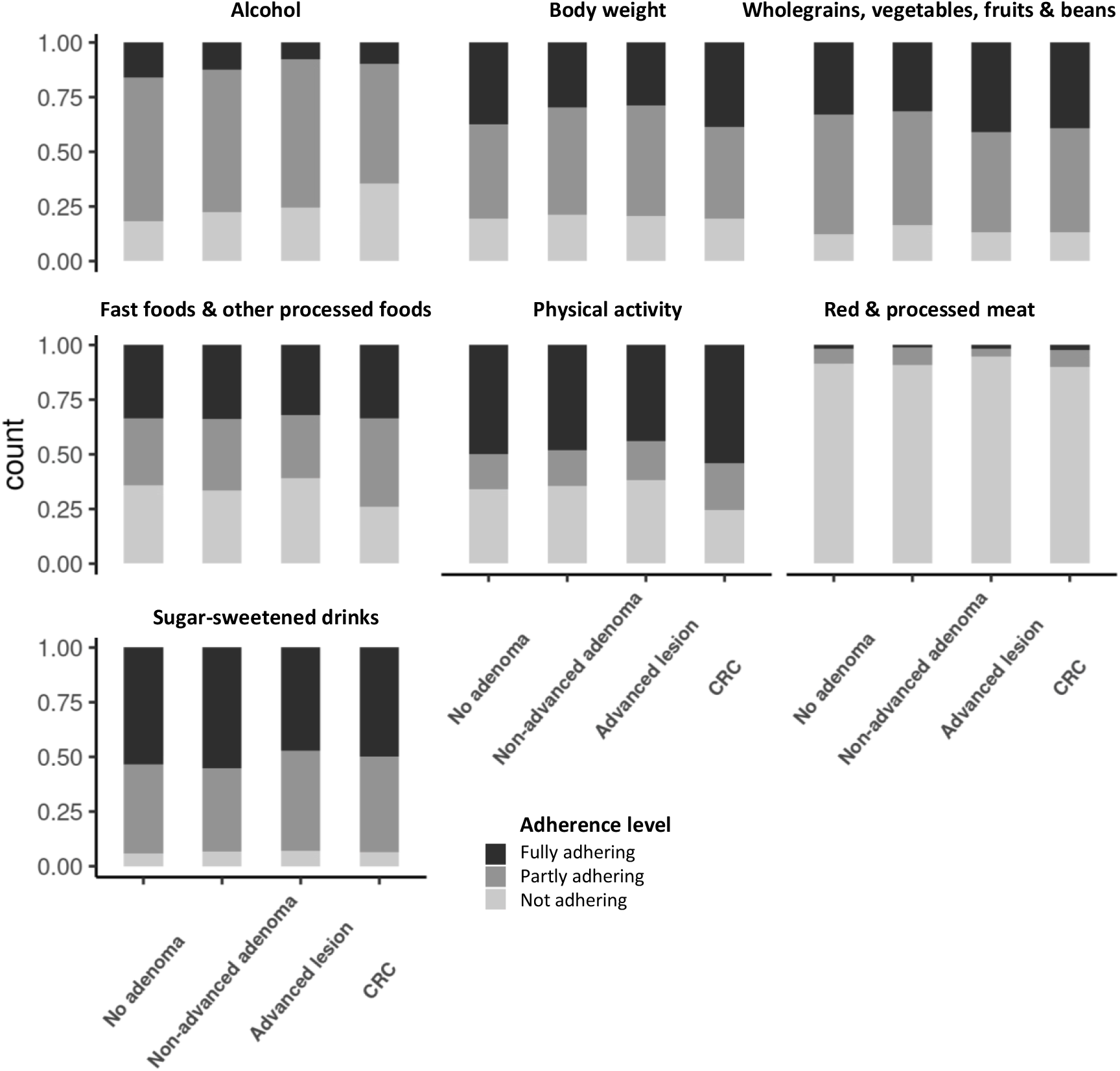
Proportion of participants who fully, partly and do not adhere to the individual Cancer Prevention Recommendations from WCRF/AICR of 2018 by stage of the carcinogenic process (n=1,914).

### The 2018 WCRF/AICR Score and stages of the carcinogenic process

Associations between adherence to the 2018 WCRF/AICR Score and presence of non-advanced adenomas, advanced lesions and CRC are shown in **Table 3**. Compared to those having the lowest adherence to the WCRF/AICR Score (≤2.5 points), participants achieving higher scores had a reduced probability of precancerous lesions, in particular advanced lesions (*p*_*trend*_ of 0.006). Compared to those having the lowest adherence, participants scoring >4.5 points had an OR (95% CI) for advanced lesions of 0.56 (0.34, 0.91). Per each point increase in the score, the probability of advanced lesions was lowered by 18% (p-value of 0.005). The inverse association was present in both sexes (although only significant in men) and irrespective of precursor lesion type (**Supplementary Figure 1**). For CRC, no associations were observed.

**Table 3.** Odds ratios (ORs) and 95% confidence intervals (CIs) for presence of non-advanced adenoma, advanced lesions and CRC relative to no adenoma by level of adherence to the 2018 WCRF/AICR Cancer Prevention Recommendations. Estimates are for the population as a whole (n=1,914) and by sex (men: n=1,057, women: n=857)^1^.

|  | Stage of the carcinogenic process |  |  |  |  |  |  |
| --- | --- | --- | --- | --- | --- | --- | --- |
|  | No adenoma<br>(n=548) | Non-advanced adenomas<br>(n=524) |  | Advanced lesions<br>(n=349) |  | CRC<br>(n=493) |  |
|  | <u>n</u> | <u>n</u> | <u>OR (95% CI)</u> | <u>n</u> | <u>OR (95% CI)</u> | <u>n</u> | <u>OR (95% CI)</u> |
| **Overall** |  |  |  |  |  |  |  |
| Points |  |  |  |  |  |  |  |
| ≤2.5 | 96 | 116 | Ref. | 84 | Ref. | 75 | Ref. |
| >2.5-3.5 | 186 | 160 | 0.75 (0.53, 1.06) | 128 | 0.81 (0.56, 1.19) | 171 | 1.18 (0.81, 1.74) |
| >3.5-4.5 | 169 | 185 | 0.91 (0.64, 1.30) | 96 | 0.65 (0.44, 0.97) | 180 | 1.37 (0.93, 2.03) |
| >4.5 | 97 | 63 | **0.59 (0.38, 0.91)** | 41 | **0.56 (0.34, 0.91)** | 67 | 0.98 (0.61, 1.56) |
| *P<sub>trend</sub>* |  |  | 0.10 |  | **0.006** |  | 0.74 |
| *Per one point increase* |  |  | 0.91 (0.80, 1.03) |  | **0.82 (0.71, 0.94)** |  | 1.04 (0.91, 1.18) |
| *P-value* |  |  | 0.13 |  | **0.005** |  | 0.60 |
| **Men** |  |  |  |  |  |  |  |
| Points |  |  |  |  |  |  |  |
| ≤2.5 | 56 | 82 | Ref. | 59 | Ref. | 47 | Ref. |
| >2.5-3.5 | 98 | 95 | 0.67 (0.42, 1.04) | 88 | 0.82 (0.51, 1.32) | 93 | 1.08 (0.66, 1.78) |
| >3.5-4.5 | 77 | 102 | 0.84 (0.53, 1.33) | 58 | 0.67 (0.40, 1.12) | 101 | 1.57 (0.95, 2.62) |
| >4.5 | 36 | 26 | **0.48 (0.26, 0.89)** | 13 | **0.36 (0.17, 0.76)** | 26 | 0.91 (0.47, 1.75) |
| *P<sub>trend</sub>* |  |  | 0.09 |  | **0.006** |  | 0.45 |
| *Per one point increase* |  |  | 0.88 (0.73, 1.05) |  | **0.80 (0.66, 0.98)** |  | 1.10 (0.91, 1.32) |
| *P-value* |  |  | 0.14 |  | **0.029** |  | 0.33 |
| **Women** |  |  |  |  |  |  |  |
| Points |  |  |  |  |  |  |  |
| ≤2.5 | 40 | 34 | Ref. | 25 | Ref. | 28 | Ref. |
| >2.5-3.5 | 88 | 65 | 0.91 (0.52, 1.61) | 40 | 0.74 (0.39, 1.39) | 78 | 1.41 (0.75, 2.63) |
| >3.5-4.5 | 92 | 83 | 1.11 (0.63, 1.94) | 38 | 0.63 (0.33, 1.19) | 79 | 1.17 (0.62, 2.20) |
| >4.5 | 61 | 37 | 0.75 (0.40, 1.42) | 28 | 0.73 (0.37, 1.47) | 41 | 1.02 (0.51, 2.03) |
| *P<sub>trend</sub>* |  |  | 0.62 |  | 0.33 |  | 0.64 |
| *Per one point increase* |  |  | 0.96 (0.80, 1.14) |  | 0.84 (0.68, 1.03) |  | 0.96 (0.80, 1.16) |
| *P-value* |  |  | 0.63 |  | 0.10 |  | 0.70 |

<sup>1</sup>Odds ratios (ORs) and 95% confidence intervals (CIs) are obtained from multinomial logistic regression analyses adjusting for the following covariates: age (continuous), sex, energy intake (continuous), smoking status (current smoker, non-smoker, missing), education level (primary school, high school, collage/university, missing) and family history of CRC (yes, no, unknown).
Abbreviations: AICR; American Institute for Cancer Research, CI; confidence interval, OR; odds ratio, Ref; reference, WCRF; World Cancer Research Fund.

### Adherence to the individual diet and lifestyle components and stages of the carcinogenic process

Associations between adherence to the individual diet and lifestyle components and stages of the carcinogenic process are shown in **Figure 3**. The strongest and most clear associations were observed for the alcohol recommendation. Compared to those not adhering to the alcohol recommendation, ORs (95% CI) for full adherence was 0.66 (0.43, 1.01), 0.39 (0.23, 0.65) and 0.32 (0.21, 0.49) for non-advanced adenoma, advanced lesions and CRC, respectively. For the precancerous lesions, full adherence to the body weight recommendation was inversely associated with lesion detection relative to non-adherence, with ORs (95% CIs) for non-advanced and advanced lesions of 0.71 (0.50, 1.00) and 0.72 (0.49, 1.06), respectively. With regard to CRC, both partial and full adherence to the physical activity recommendation and the recommendation concerning “fast foods” and other processed foods were positively associated with having a cancer diagnosis (ORs (95% CIs) of 1.49 (1.12, 1.98) and 1.44 (1.05, 1.96) for full adherence to the physical activity and ‘fast foods’ recommendation, respectively).

**Figure 3.**
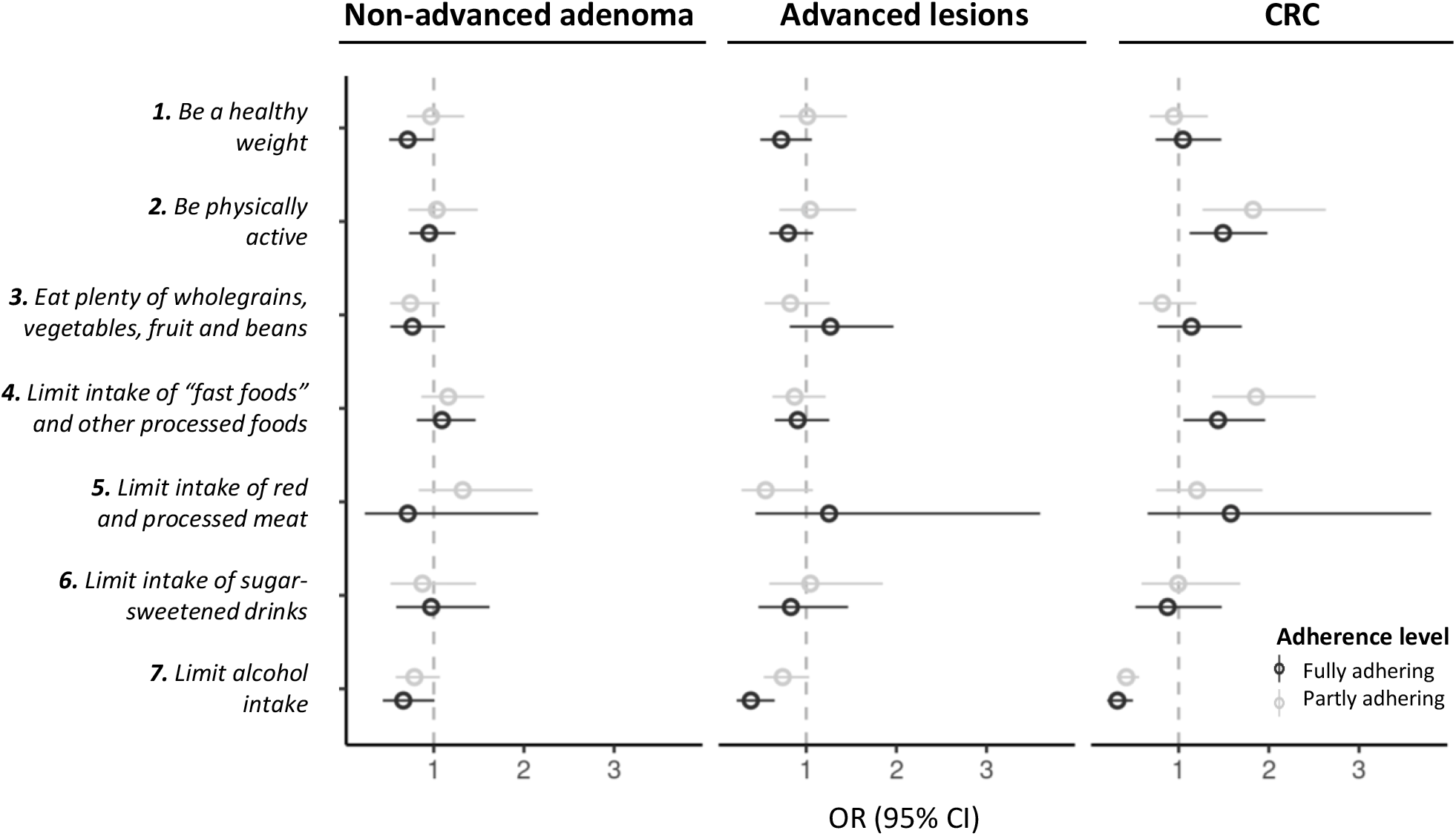
Odds ratios (ORs) and 95% confidence intervals (CIs) for presence of non-advanced adenoma, advanced lesions and CRC relative to no adenoma by adherence to the individual 2018 WCRF/AICR Cancer Prevention Recommendations in the study population as a whole (n=1,914). Non-adherence (0 points) is treated as the reference category. Analyses are adjusted for age (continuous) and sex.

### Sensitivity analyses

Neither timing of cancer care nor disease severity modified the relationship between the 2018 WCRF/AICR Score and presence of a cancer diagnosis (**Supplementary Figure 2**). Restricting the main analyses to those recruited from the CRCbiome study (1,484 (78%), of which 63 (4%) were diagnosed with CRC), did not change the interpretation of the findings (Supplementary Table 2).

## Discussion

In this large cross-sectional investigation among participants at different stages of CRC development, adhering to the 2018 WCRF/AICR Score was inversely associated with presence of precancerous lesions, in particularly advanced colorectal lesions, but not CRC. Adherence to the alcohol recommendation was the single factor most strongly inversely associated with CRC development, associations being observed across all carcinogenic stages. For the remaining recommendations, results were less coherent, suggesting that adherence to the recommendations as a whole may be more important for cancer prevention than adherence to each factor separately.

Various studies have investigated the association of adherence to either the 2007 (19–21) or 2018 (22–25) edition of the cancer prevention recommendations from WCRF/AICR and risk of CRC. However, to the best of our knowledge, no study before the present has assessed the associations across the entire spectrum of colorectal carcinogenesis. Using this approach, we demonstrate that adherence to the 2018 WCRF/AICR cancer prevention recommendations was strongly inversely associated with detection of precancerous lesions, in particular the high-risk lesions, in a dose-response manner. The lowest probability of lesion detection was seen among those adhering to just above half (>4.5/7 points) or more of the recommendations. The inverse associations were observed irrespective of histopathological subtype and among both sexes. In a comparable study, Erben, *et al*. (26). investigated the associations of a healthy diet and lifestyle score with colorectal lesions, also representing the entire spectrum of CRC development. In that cross-sectional investigation, including more than 13,000 German screening participants, strong inverse associations were observed with presence of hyperplastic polyps, non-advanced adenomas and advanced colorectal neoplasms, the latter group consisting mostly of advanced adenomas (>90%). Inverse associations between a healthy lifestyle pattern and precancerous colorectal lesions have also been observed in the main BCSN pilot population (27–29). Together, these findings support the importance of adhering to diet and lifestyle recommendations to prevent early-stage colorectal carcinogenesis.

There are several potential reasons for the unexpected lack of association between adherence to the cancer prevention recommendations and a diagnosis of CRC in the present population. First, the cross-sectional design makes the study prone to reverse causality. Receiving a lifestyle-related disease may have given the participants an incentive to improve their overall lifestyle to favor prognosis. It is also possible that the occurrence of a gastrointestinal tumor or the subsequent treatment have led to unvoluntary changes in lifestyle of relevance to the score (e.g. a reduction in BMI due to disease-related malnutrition (30)). In a prospective study among CRC survivors (31), van Zutphen, *et al*. investigated the adherence to the standardized 2018 WCRF/AICR Score from time of diagnosis to 6 and 24 months follow-up. Although only marginal changes were observed in the overall lifestyle, the majority of participants (92%) changed adherence to at least one recommendation, half undergoing simultaneous changes leading to improvement in one component and a deterioration of another. It is possible that such fluctuations may have introduced noise in our analyses, given the difference in time for which the diet and lifestyle registration was performed (from weeks prior to screening colonoscopy to almost a year following CRC surgery). However, neither time of cancer care, nor the progressive nature of the disease – as a main determinant of treatment complexity - influenced the result. It should be noted though, that the separation of CRC patients into smaller subgroups, greatly reduced the sample size, limiting statistical power.

In the present study, adherence to the alcohol recommendation was the single factor most strongly inversely associated with CRC development, being associated with all stages of colorectal carcinogenesis. That higher alcohol intake is linked to carcinogenic development is supported by a meta-analysis on adenoma risk (32), as well the latest expert report from WCRF/AICR on CRC (2). In a separate investigation of the Global Burden of Disease Project focusing on the alcohol-attributed cancer burden (33), it was estimated that harmful alcohol use contributed to ∼4% of all incident cancers and about one in ten cases originating from the colon or rectum. Increasing the public’s awareness of the importance of adhering to the alcohol recommendation is of outmost importance to lower the cancer burden attributable to this risk factor.

Except for adherence to the alcohol recommendation, associations between the other recommendations and stages of the carcinogenic process were less clear. This could suggest that adhering to multiple recommendations in combination - as an integrated package of lifestyle behaviors – is more important for CRC prevention than adherence to individual factors alone. Indeed, the importance of taking a holistic approach to cancer prevention represents one of the major shifts in focus in the cancer prevention recommendations of 2018 compared to earlier versions.

It is also possible that there are better ways of operationalizing the recommendations. For instance, we were not able to show an association between adherence to the red and processed meat recommendation and presence of any colorectal lesions, although evidence linking these food items to CRC development is considered strong (2,34), and also shown in the CRCbiome population (35). Only partial adherence to the meat recommendation was borderline significantly associated with advanced lesions. This could suggest that the cut points to achieve full score, particularly those for processed meat (<3 g/day, fulfilled by only 2% of participants) were unnecessarily strict. A recent comparative analysis of 18 dietary patterns and risk of CRC (25) suggests a potential for further refining the 2018 WCRF/AICR recommendations by making use of already available dietary patterns (e.g. those reflecting hyperinsulinemia, hypertension, chronic inflammation and a Western dietary pattern).

A major strength of the present study is the pooling of data from two large cancer studies, resulting in a unique study sample reflecting the entire spectrum of colorectal carcinogenesis. The high number of clinically verified precancerous and cancerous lesions increased the power and precision of our association analyses. A further strength is the use of a standardized scoring system for measuring adherence to the 2018/AICR cancer prevention recommendations, enabling cross-study comparisons (3,4). The access to comprehensive high-quality data on diet and lifestyle, allowed a thorough evaluation of adherence to each recommendation and the operationalization was carried out by two registered dietitians.

The main limitation of the study is the cross-sectional design, implications of which have been thoroughly discussed above. The skewed contribution of CRC cases (∼90% being recruited from CRC-NORDIET) may also have introduced bias. Although more or less similar assessment tools were used to characterize participants’ body weight, physical activity and diet in the two studies, differences in study design (observational *vs*. experimental study) and associated barriers for participation, could limit the comparability between the studies. There is a general concern that participants of clinical trials reflect the healthiest, most educated subpart of the population (38), and threats to external validity have been documented in several cancer clinical trials (39–41). It is possible that differences in selection of participants into the two studies distorted the associations for CRC in the present study. Lastly, the findings could be limited by the use of self-reported data for construction of the score. However, the questionnaires used have been validated for the majority of components included in the score (8,9,42–45), and mostly shown to produce acceptable results.

To conclude, in this unique sample of participants reflecting the entire spectrum of colorectal carcinogenesis, high adherence to the 2018 WCRF/AICR cancer prevention recommendations was inversely associated with presence of precancerous lesions, in particular advanced colorectal lesions, but not CRC. Except for adherence to the alcohol recommendation, where strong inverse associations were observed across all carcinogenic stages, associations for adherence to the other recommendations were less consistent. For early-stage colorectal carcinogenesis, the largest preventive effects could likely be achieved by adhering to multiple cancer prevention recommendations in combination, when followed together, promote a healthy pattern of diet and physical activity conducive to the prevention of cancer.

## Data Availability

The data generated in the two studies are not publicly available due to the principles and conditions set out in articles 6 [1] (e) and 9 [2] (j) of the General Data Protection Regulation (GDPR), but are available upon reasonable request from the principles investigators: CRCbiome; Paula Berstad and Trine B. Rounge, CRC-NORDIET; Rune Blomhoff.

## Acknowledgements

We would like to thank the devoted healthcare personal at Bærum and Moss hospitals, Oslo University Hospital, Ullevål and Akershus University Hospital for their important contribution to the CRCbiome and CRC-NORDIET study. We would also like to thank the administrative personal, technicians and students that have played crucial roles in the data collection and processing of the two studies. Lastly, we would like to thank each and every participant taking part; without you, the conduction of the two studies would not have been possible.

## Authors’ contributions

Conception and design: ASK, HBH, PB and RB

Development of methodology: ASK, ARA, HBH, PB and RB

Acquisition of data (provided animals, acquired and managed patients, provided facilities, etc.): ASK, ARA, HBH, SKB, IP, GW, SS, TBR, RB and PB

Analysis and interpretation of data (e.g., statistical analysis, biostatistics, computational analysis): ASK, ARA, HBH, PB and RB

Writing, review, and/or revision of the manuscript: ASK, ARA, HBH, MK, AMWJ, AH, SKB, IP, GW, SS, TBR, RB and PB

Administrative, technical, or material support (i.e., reporting or organizing data, constructing databases): ASK, ARA, HBH and AMWJ

Study supervision: ASK, HBH, PB and RB

## List of abbreviations

AICR: American Institute for Cancer Research
BCSN: Bowel Cancer Screening in Norway
CRC: Colorectal cancer
FFQ: Food frequency questionnaire
FIT: Fecal immunochemical test (FIT)
ICD: International classification of diseases
LDQ: Lifestyle- and demograpxhics questionnaire
KBS: Kostberegningssystem (“Dietary calculation system”)
NCT: National clinical trial
WCRF: World Cancer Research Fund

