## Supplementary material for "Associations of the 2018 World Cancer Research Fund/American Institute of Cancer Research (WCRF/AICR) Cancer Prevention Recommendations with Stages of Colorectal Carcinogenesis"

### **Supplementary Data**

**Supplementary Table 1.** The 2018 WCRF/AICR Cancer Prevention Recommendations with suggested^1^ and applied operationalization.

| **2018 WCRF/AICR Recommendations with suggested operationalization** | | | **Operationalization in the current study** | |
| --- | --- | --- | --- | --- |
| 2018 WCRF/AICR recommendations | Recommend operationalization of recommendations with suggested scoring | Suggested scoring | Applied scoring | Comments and considerations |
| **1. Be a healthy weight** | **BMI (kg/m^2^):**  18.5–24.9  25–29.9  <18.5 or ≥30  **Waist circumference (cm):**  M: <94 / W: <80  M: 94–<102 / W: 80–<88  M: ≥102 / W: ≥88 | 0.5  0.25  0  0.5  0.25  0 | 1.0  0.5  0  -  -  - | Only BMI available for both studies  Calculated from self-reported body weight (kg) and height (m^2^) |
| **2. Be physically active** | **Total moderate-vigorous physical activity (min/wk):**  ≥150  75–<150  <75 | 1  0.5  0 | 1  0.5  0 | Calculated as the sum of self-reported moderate (min/wk) and vigorous (min/wk) physical activity, the latter weighted by a factor of two to best reflect the recommendation |
| **3. Eat a diet rich in wholegrains, vegetables, fruit and beans** | **Fruits and vegetables (g/day):**  ≥400  200–<400  <200  **Total fiber (g/day):**  ≥30  15–<30  <15 (0) | 0.5  0.25  0  0.5  0.25  0 | 0.5  0.25  0  0.5  0.25  0 | Fruits and vegetables included all fresh, frozen and conserved fruits, berries and vegetables.  For the following items, a prespecified proportion was included to account for other ingredients in the product:   - Jam and marmalade (50%) - Vegetable dishes (50%)   The food item ‘vegetable soup’ was left out to lower the chances of overestimation due to the expected high contribution of dried soups.  Juice and juice concentrate were not included. Legumes were also left out. |
| **4. Limit consumption of “fast foods” and other processed foods high in fat, starches or sugars** | **Percent of total kcal from ultra-processed foods (aUPFs):**  Tertile 1  Tertile 2  Tertile 3 | 1  0.5  0 | 1  0.5  0 | The aUPF variable was constructed based on the NOVA classification system(36). Food items already included in other components of the score (e.g. sugar-sweetened drinks and red and processed meats) were left out to avoid double penalization.  The following items were defined as aUPFs: White bread, tortilla, chapatti and related products, sandwich biscuits, breakfast cereals with added sugar, cakes, desserts, ice cream, sorbet, chocolates, sweets, snacks, processed products of milk or cream such as vanilla sauce, honey/syrup (50% included), jam/marmalade and other spreads with added sugar, artificial sweeteners, margarine and mixed products of margarine and butter (50% included), mayonnaise, french fries, mashed potatoes, vegetable soups, vegetable products, fish products, sauces, dry soups, bouillon powder/cubes, artificially sweetened lemonade and soda, milk substitutes and liquor  The aUPF variable was constructed by adding up the listed items in kcal/day. |
| **5. Limit consumption of red and processed meat** | **Total red meat (g/wk) and processed meat (g/wk):**  Red meat <500 and processed meat <21  Red meat <500 and processed meat 21–<100  Red meat >500 or processed meat ≥100 | 1  0.5  0 | 1  0.5  0 | Red meat included all non-white meat, except wild game meat. Both processed and non-processed red meat were included.  Processed meat included all processed meat products, irrespective of animal origin. |
| **6. Limit consumption of sugar-sweetened drinks** | **Total sugar-sweetened drinks (g/day):**  0  >0–≤250  >250 | 1  0.5  0 | 1  0.5  0 | Sugar-sweetened drinks included lemonade, soda and milk with added sugar, juice concentrate and mixed drinks. |
| **7. Limit alcohol consumption** | **Total ethanol (g/day):**  0  M: >0–≤28 (2 drinks) / W: ≤14 (1 drink)  M: >28 (2 drinks) / W: >14 (1 drink) (0) | 1  0.5  0 | 1  0.5  0 |  |
| **8. For mothers: breastfeed your baby, if you can (optional)** | **Exclusively breastfed over lifetime for a total of:**  6+ months  >0–<6 months  Never | 1  0.5  0 | -  -  - | Data not available |

*^1^Shams-White, et al. Operationalizing the 2018 World Cancer Research Fund/American Institute for Cancer Research (WCRF/AICR) Cancer Prevention Recommendations: A Standardized Scoring System. Nutrients (2019).*

| **Supplementary Table 2.** Odds ratios (ORs) and 95% confidence intervals (CIs) for presence of non-advanced adenoma, advanced lesions and CRC relative to no adenoma by adherence to the 2018 WCRF/AICR Cancer Prevention Recommendations. Only the CRCbiome participants are included in the analyses (n=1,484)^1^. | | | | | |
| --- | --- | --- | --- | --- | --- |
|  | ≤2.5 points (n=306) | >2.5-3.5 points (n=493) | >3.5-4.5 points (n=473) | >4.5 points (n=212) | *p trend* |
| **Non-advanced adenoma** |  |  |  |  |  |
| No. of events | 116 | 160 | 185 | 63 |  |
| *Model 1* | Ref. | 0.71 (0.50, 1.00) | 0.87 (0.62, 1.23) | **0.53 (0.35, 0.80)** |  |
| *Model 2* | Ref. | 0.72 (0.51, 1.02) | 0.91 (0.64, 1.29) | **0.58 (0.38, 0.88)** | 0.10 |
| **Advanced lesions** |  |  |  |  |  |
| No. of events | 84 | 128 | 96 | 41 |  |
| *Model 1* | Ref. | 0.78 (0.54, 1.13) | 0.63 (0.43, 0.92) | **0.48 (0.30, 0.76)** |  |
| *Model 2* | Ref. | 0.81 (0.56, 1.17) | **0.67 (0.45, 0.99)** | **0.56 (0.35, 0.90)** | **0.007** |
| **CRC** |  |  |  |  |  |
| No. of events | 10 | 19 | 23 | 11 |  |
| *Model 1* | Ref. | 0.97 (0.44, 2.18) | 1.27 (0.58, 2.78) | 1.08 (0.44, 2.65) |  |
| *Model 2* | Ref. | 1.00 (0.45, 2.23) | 1.32 (0.60, 2.90) | 1.15 (0.46, 2.87) | 0.54 |

*^1^Odds ratios (ORs) and 95% confidence intervals (CIs) are obtained using multinomial logistic regression models adjusted for the following covariates: Model 1: age (continuous) (n=1,484), model 2: age (continuous), sex and energy intake (continuous) (n=1,484).*

*Abbreviations: AICR; American Institute for Cancer Research, CI; confidence interval, CRC; Colorectal cancer, OR; odds ratio, Ref; reference, WCRF; World Cancer Research Fund.*

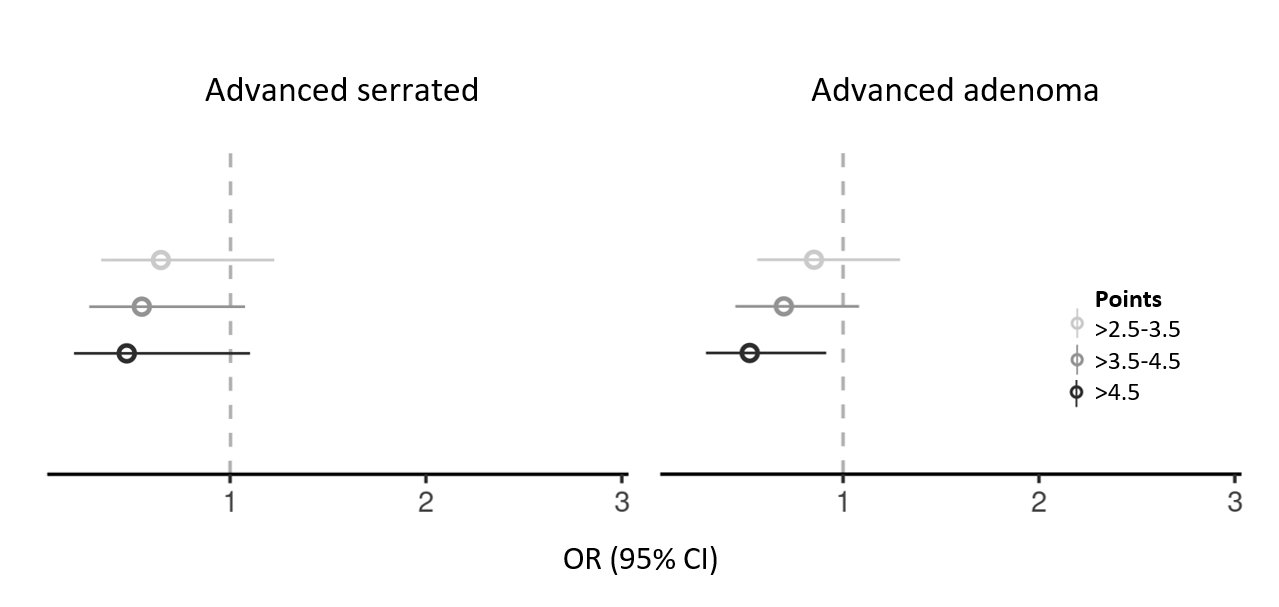

**Supplementary Figure 1**. Odds ratios (ORs) and 95% confidence intervals (CIs) for presence of the two main precursor lesions advanced adenoma and advanced serrated lesion relative to no adenoma by adherence to the 2018 WCRF/AICR Cancer Prevention Recommendations. Effect estimates are derived from a multinomial logistic regression analysis, including the following clinical groups: No adenoma (n=548), non-advanced adenoma (n=524), advanced serrated lesion (n=74), advanced adenoma (n=238), mixed lesions (n=37) and CRC (n=493). The low adherence group (≤ 2.5 points) is treated as the reference category. Analyses are adjusted for age (continuous) and sex.

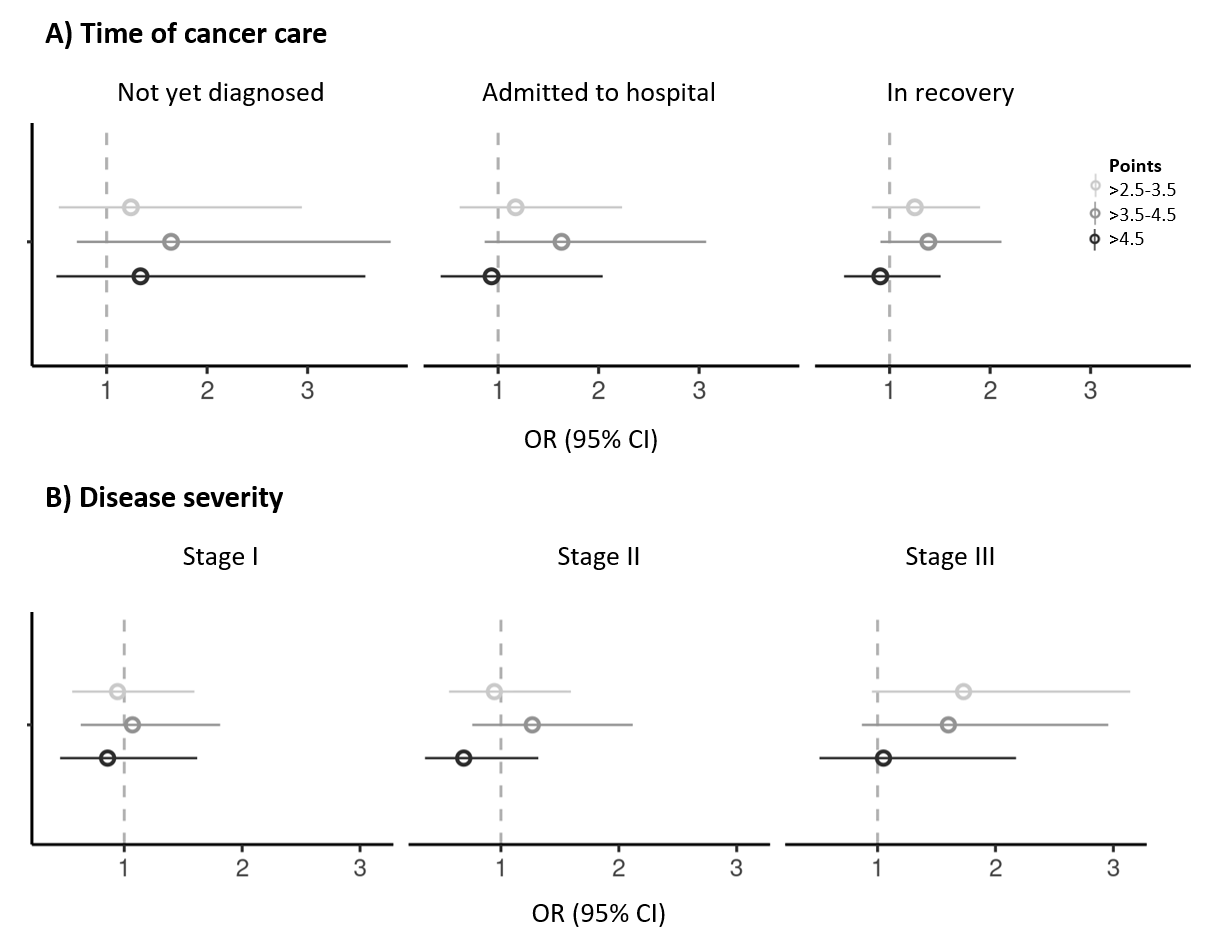

**Supplementary Figure 2**. Odds ratios (ORs) and 95% confidence intervals (CIs) for presence of CRC by time of FFQ completion (**A**) and TNM stage (**B**) relative to no adenoma by adherence to the 2018 WCRF/AICR Cancer Prevention Recommendations. Effect estimates are derived from multinomial logistic regression analyses, including the following clinical groups: No adenoma (n=548), non-advanced adenoma (n=524), advanced lesion (n=349) and CRC (A: Not yet diagnosed (n=60), admitted to hospital (n=107) and in recovery (n=320), B: Stage I (n=151), stage II (n=160) and stage III (n=137)). The low adherence group (≤ 2.5 points) is treated as the reference category. Analyses are adjusted for age (continuous) and sex.
